# Kinetics and Isotype Assessment of Antibodies Targeting the Spike Protein Receptor Binding Domain of SARS-CoV-2 In COVID-19 Patients as a function of Age and Biological Sex

**DOI:** 10.1101/2020.07.15.20154443

**Authors:** Nancy R. Graham, Annalis N. Whitaker, Camilla A. Strother, Ashley K. Miles, Dore Grier, Benjamin D. McElvany, Emily A. Bruce, Matthew E. Poynter, Kristen K. Pierce, Beth D. Kirkpatrick, Renee D. Stapleton, Gary An, Jason W. Botten, Jessica W. Crothers, Sean A. Diehl

**Affiliations:** Department of Microbiology and Molecular Genetics, Larner College of Medicine University of Vermont, Burlington, VT, 05405, USA; Vaccine Testing Center, Larner College of Medicine University of Vermont, Burlington, VT, 05405, USA; Department of Medicine-Immunobiology, Larner College of Medicine University of Vermont, Burlington, VT, 05405, USA; Cellular, Molecular, and Biomedical Sciences Graduate Program, Larner College of Medicine University of Vermont, Burlington, VT, 05405, USA; Vermont Center for Immunology and Infectious Disease, Larner College of Medicine University of Vermont, Burlington, VT, 05405, USA; Department of Pathology and Laboratory Medicine, Larner College of Medicine University of Vermont, Burlington, VT, 05405, USA; Vermont Lung Center, Larner College of Medicine University of Vermont, Burlington, VT, 05405, USA; Department of Medicine-Pulmonary and Critical Care, Larner College of Medicine University of Vermont, Burlington, VT, 05405, USA; Medicine-Infectious Disease, Larner College of Medicine University of Vermont, Burlington, VT, 05405, USA; Department of Surgery, Larner College of Medicine University of Vermont, Burlington, VT, 05405, USA; Translational Global Infectious Disease Research Center, Larner College of Medicine University of Vermont, Burlington, VT, 05405, USA

**Keywords:** COVID-19, SARS-CoV-2, spike protein, Receptor-binding domain, Coronavirus, Serology, Humoral immune response, Neutralizing antibody, Isotypes

## Abstract

SARS-CoV-2 is the newly emerged virus responsible for the global COVID-19 pandemic. There is an incomplete understanding of the host humoral immune response to SARS-CoV-2 during acute infection. Host factors such as age and sex as well the kinetics and functionality of antibody responses are important factors to consider as vaccine development proceeds. The receptor-binding domain of the CoV spike (RBD-S) protein is important in host cell recognition and infection and antibodies targeting this domain are often neutralizing. In a cross-sectional study of anti-RBD-S antibodies in COVID-19 patients we found equivalent levels in male and female patients and no age-related deficiencies even out to 93 years of age. The anti-RBD-S response was evident as little as 6 days after onset of symptoms and for at least 5 weeks after symptom onset. Anti-RBD-S IgG, IgM, and IgA responses were simultaneously induced within 10 days after onset, but isotype-specific kinetics differed such that anti-RBD-S IgG was most sustained over a 5-week period. The kinetics and magnitude of neutralizing antibody formation strongly correlated with that seen for anti-RBD-S antibodies. Our results suggest age- and sex-related disparities in COVID-19 fatalities are not explained by anti-RBD-S responses. The multi-isotype anti-RBD-S response induced by live virus infection could serve as a potential marker by which to monitor vaccine-induced responses.

## Introduction

Human pathogenic coronaviruses (CoV) such as severe acute respiratory syndrome (SARS)-CoV-1, middle east respiratory syndrome (MERS)-CoV, and SARS-CoV-2 (all β-CoVs) have resulted from zoonoses and utilize cellular receptors to bind and access host cells for productive infection (1–3). CoV spike (S) proteins are large (>200 kDa) glycosylated trimeric structures that protrude from viral particles and enable binding of CoV to cellular receptors. SARS-CoV-2 interacts with angiotensin converting enzyme-2 (ACE2) via a flexible receptor-binding domain (RBD) located on the distal tip of the S protein (4–7). After binding, several proteases act upon S, priming it to adopt large conformational shifts that facilitate entry into host cells(8). First the S1 domain (which contains RBD) is cleaved from the C-terminal S2 domain. For SARS-CoV-2 this process may involve furin in the host cell membrane due to a novel furin-recognition site in the S1/S2 region (9–11). The S2 domain is further processed by other serine and cysteine-proteases such as trypsin, cathepsin, and TMPRSS2 to facilitate viral entry into the host cell (4, 12).

Neutralizing antibodies to SARS CoV-1 have been isolated and were found to target RBD-S (13). One of these mAbs CR3022 was also found to bind SARS-CoV-2 RBD-S(14). At the polyclonal level, the quantity of anti-RBD S IgG antibodies against SARS-CoV-2 correlate well with neutralizing activity(15–18). Cross-neutralization amongst SARS viruses by RBD-S-targeting antibodies can occur (18–21). However, sequence homology for RBD-S is low for non-SARS β-CoVs (such as MERS) and for α-CoVs such as NL63, OC43, 229E, and HKU1(16, 17). For these reasons serology for SARS-CoV-2 RBD-S is being used to help identify recovered COVID-19 patients as plasma donors for passive immunotherapy (22).

There are several risk factors for COVID-19 mortality but whether two of these – age and biological sex – are associated with the SARS-CoV-2 RBD-S immune response has to our knowledge not been addressed in the peer-reviewed literature. Furthermore, most serology studies have been done in the setting of severe COVID-19 disease and, save for one study (17), without the benefit of detailed kinetics. Herein we tracked the kinetics and magnitude of neutralizing and anti-SARS-CoV-2 S and RBD-S antibodies in a cross-sectional cohort of PCR-confirmed COVID-19 patients.

## Results and Discussion

We chose a two-step ELISA–based RBD-S-focused approach to serology in our study population. Reagents and pre-print protocols were available in mid-March 2020, which indicated that RBD-S screening and full-S confirmation could identify specific and functional antibodies and be quickly operationalized. Using the established protocol (23) we confirmed the expected protein size of mammalian-expressed RBD-S (**Figure 1A**) and trimerized spike (**Figure 1B**) produced from DNA plasmids (gift from Florian Krammer, Mt Sinai School of Medicine). RBD-S antibodies were specific and correlated with neutralization (15), findings that have been validated using similar RBD-S-focused assays(16, 17). We confirmed RBD-S and S protein conformation by binding of CR3022 human IgG1 (**Figure 1C, D**). CR3022 was isolated as a SARS-S1 domain-binding single chain antibody fragment by phage display and is neutralizing as an IgG1(13). CR3022 binds adjacent to RBD-S in trimeric S of SARS-CoV-2 in a glycosylation-sensitive manner(14). Mammalian expression of appropriate size proteins and recognition by CR3022 together confirm that our protein preparations exhibited the expected characteristics.

**Figure 1.**
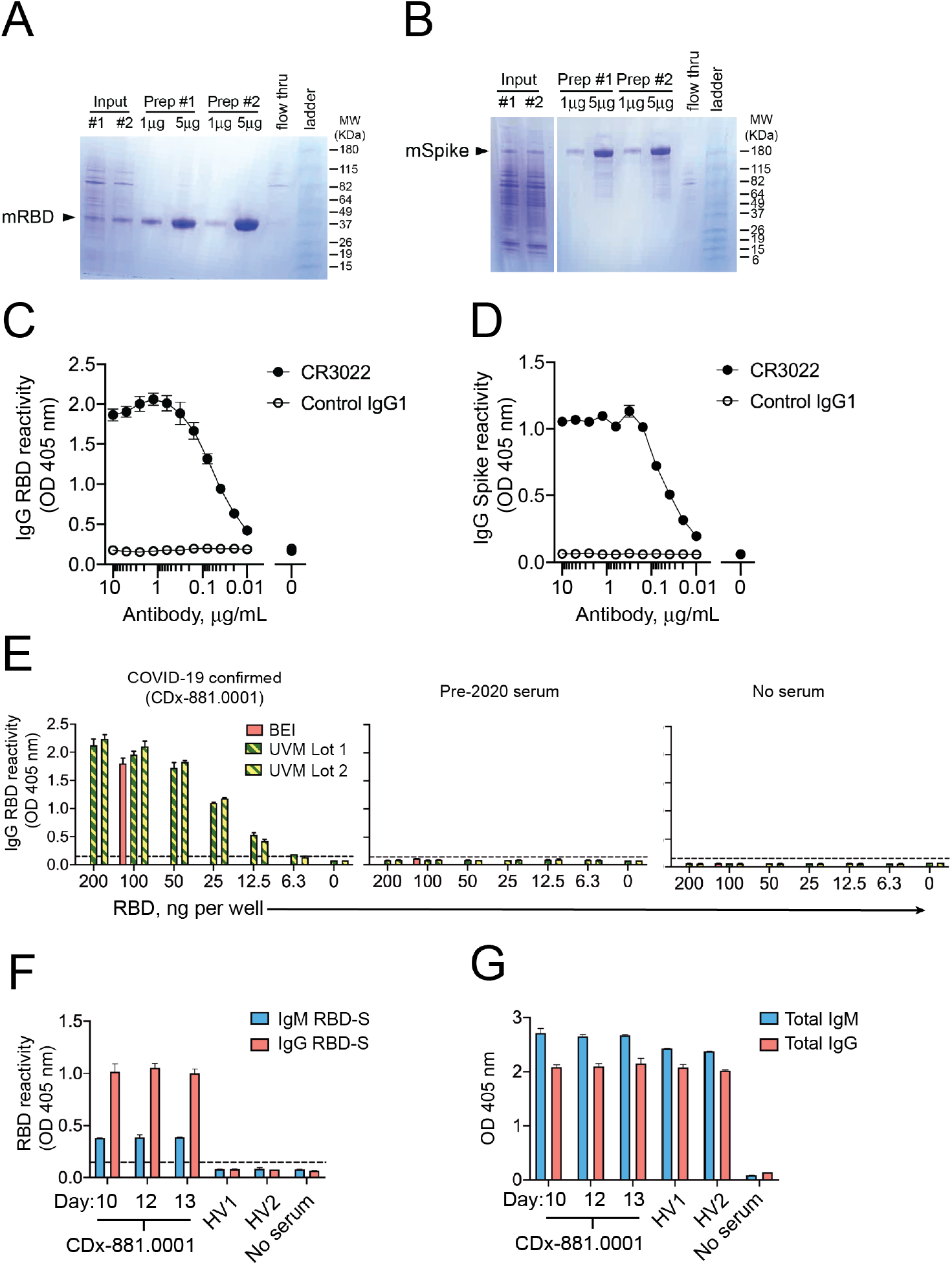
Validation of SARS-CoV-2 RBD-S and spike antigens in COVID-19 samples. Reducing SDS-PAGE analysis of (**A**) RBD-S and (**B**) trimeric spike purified from transiently transfected mammalian HEK293 cells. (**C**) Binding of CR3022 IgG1 mAb to SARS-CoV-2 RBD-S and (**D**) trimerized spike. The anti-dengue virus 1M7 mAb (35) was used as a control (**E**) Detection of serum IgG from a COVID-19 patient (left), but not from pre-2020 serum (center) or no serum control (right). (**F**) Detection of IgM and IgG to RBD-S in serial serum samples from COVID-19 patient and not in pre-2020 healthy volunteer sera (all sera diluted 1:50 and for COVID-19 patient, day after onset is shown in label). (**G**) Total IgM and IgG reactivity in a 1:50 dilution of serum from panel F.

We first piloted our antigen preps for the RBD-S IgG screening assay using serum samples from a PCR-confirmed severe COVID-19 patient (defined as admission to the Intensive Care Unit, ICU) who was admitted to the hospital 10 days following symptom onset and based on an early report suggesting that SARS-CoV-2 could trigger antibody responses in this timeframe (24). We compared IgG reactivity in this sample to decreasing amounts of our RBD-S antigen preparations against a fixed, recommended amount of commercially produced RBD-S protein derived from the protocol we used (23). We found that a wide range of locally produced RBD-S antigen yielded IgG reactivity equivalent to 100 ng of commercial antigen in an acute serum sample from this COVID-19-positive patient (**Figure 1E**). No signal was observed in a pre-2019 serum sample or in the absence of serum (**Figure 1E**). Using the standard 100 ng amount hereafter, we found that RBD-S–binding IgM and IgG were present at 10-13 days after symptom onset. We did not detect any RBD-S-binding in healthy pre-2019 sera (**Figure 1F**), in agreement with extensive testing of this assay in pre-COVID-19 serum performed elsewhere (15). Due to different secondary antibodies for IgM and IgG detection we cannot conclude whether absolute levels of RBD-S IgG were higher than RBD-S IgM. Total IgG and IgM were readily detected in both COVID-19 and in healthy non-COVID-19 serum (**Figure 1G)**.

For a cross-sectional COVID-19 serological survey we collected serum samples from 32 patients that tested COVID-19 positive by nasopharyngeal swab RT-qPCR testing. All patients had been admitted to the hospital and 13/32 (40%) were admitted to the ICU. Twenty-five patients were subsequently discharged and 7 died. One to five serum samples were collected from each patient with the first sample being taken within approximately 9 days after diagnosis, in which diagnosis occurred around 5 days after symptom onset (**Table 1**). There was a 53%:47% male: female distribution and patients were on average 68 ± 14 years of age (range 30–93 years) (**Table 1**).

**Table 1.**

| **COVID-19**<br>**subjects** | **Male/**<br>**Female** | **AGE ± S.D.**<br>**[Range]** | **Days from**<br>**symptoms to**<br>**Dx** | **Days between**<br>**Dx and 1<sup>st</sup>**<br>**serum** |
| --- | --- | --- | --- | --- |
| Swab PCR+ (n =<br>32) | 17/15 | 68 ± 14 [30-93] | 5.4 ± 4.7 [0-14] | 8.6 ± 7.5 [0-35] |

A male bias in COVID-19 mortality was reported early during the pandemic (25–27) and has been confirmed worldwide in a recent meta-analysis (28). One of the hypotheses to explain this is differences in adaptive immunity between males and females. Although the mean serum RBD-S IgG reactivity level appeared higher in male samples (O.D. = 1.8, n = 40) versus female samples (O.D. = 1.0, n = 37) this difference was not significant and the same maximum reactivity values were found in males and females (**Figure 2A**).

**Figure 2.**
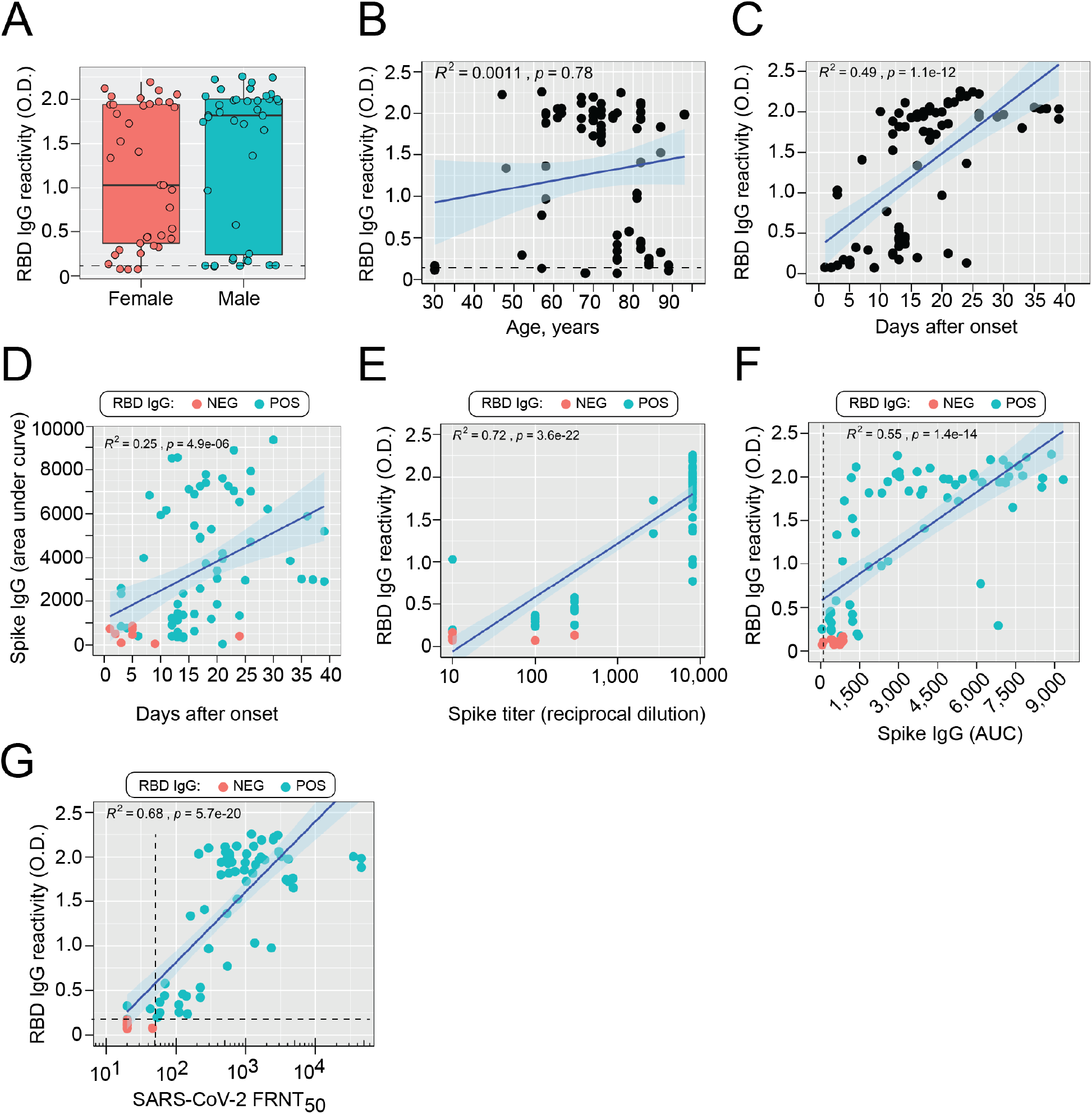
IgG responses to SARS-CoV-2 RBD-S and spike. **(A)** Comparison of RBD-S IgG reactivity (OD 405nm) levels in samples from male or female patients (*P* = 0.18, student’s t-test) Note that multiple samples came from some patients. Boxplots show the 25-75^th^ percentiles, with median as horizontal line and whiskers as 95% confidence level and all individual samples. (**B**) RBD-S IgG reactivity was assessed as a function of age (**C**) or days after symptom onset. (**D**) Spike reactivity is expressed as area under the curve over six threefold serial dilutions (1:100 – 1:8,100) and plotted against days of symptoms. (**E**) Spike IgG endpoint titer or (**F**) AUC is plotted against RBD-IgG reactivity. (**G**) SARS-CoV-2 microneutralization titers are plotted against RBD-S IgG reactivity. Cutoff values (dashed line) are shown. Spearman’s Rho coefficient (*R*^*2*^), 95% confidence interval (shading), and *P*-value are shown for B-G.

Although not absolute, it appears that irrespective of comorbidities, there is a higher risk of COVID-19 mortality and morbidity in older individuals (60 years of age and over) (29–31). We therefore assessed RBD-S IgG antibodies by age. There was a broad range of RBD-S IgG responses that did not differ as a function of age as assessed by correlation analysis (*R*^*2*^ < 0.01, **Figure 2B**). Notably, one of the highest RBD-S IgG responses was from a 93-year old patient. A serum sample from a 30-year old COVID-19 patient was negative for RBD-S IgG, but this sample was taken just three days after symptom onset, which may be too early for induction of robust IgG responses. Taken together, we did not find evidence of biological sex- or age-related deficiencies in RBD-S IgG responses in COVID-19 patients.

RDB-S-reactive serum IgG was detected in 5 of 12 (42%) samples that were taken within 10 days of symptom onset (**Figure 2C**). After day 10 of symptoms, 98% of samples were positive for RBD IgG (**Figure 2C**). There were small variations in positive threshold for RBD by assay date (Figure S1). We therefore confirmed each sample (whether RBD-positive or not) with an endpoint titration and area under the curve calculation for reactivity against the full spike ectodomain trimer (15). Samples that were RBD-S-negative were also low for spike total reactivity (AUC) and titer (**Figures 2D, E**). Furthermore, we found a very strong correlation between RBD and spike IgG (**Figure 2F**). The low level of spike reactivity in RBD-negative samples could indicate a baseline cross-reactivity against other human coronaviruses (32). S cross-reactivity would presumably occur in regions outside the RBD given the low conservation of SARS-CoV-2 RBD compared to other human CoVs with the exception of SARS-CoV-1 (16). Nonetheless, we found a strong correlation between RDB-S IgG and microneutralization titers (**Figure 2G**), confirming the utility of RBD-S serology for estimation of functional neutralizing antibodies in agreement with other studies (15–17).

In the patient-specific RBD IgG data (**Figure S2A**) we found several patterns: initial seroconversion (e.g. patients 0003, and 0017), rapid increases (e.g. patients 0005, 0006, 0009, 0011, 0020, occurring between days 10-20), and plateaued responses (e.g. patients 0012 and 0021, occurring mainly after day 20). These responses were concordant with temporal patient-specific S IgG titers (**Figure S2B**). Anti-S titers in patients with a negative RBD-S test were generally low and in RBD-positive samples, followed the same trends as RBD-reactivity, providing further confirmation of robust serological responses to SARS-CoV-2 during acute COVID-19. At the patient level, neutralizing activity was observed after as few as five days after symptom onset and throughout the study period and was predominantly found in those samples with positive RBD-S IgG (**Figure S3**).

To assess antibody isotype dynamics during acute SARS-CoV-2 we followed RBD-S and full spike-specific IgM and IgA levels in the same samples for which RBD-S and spike IgG was determined. At the patient level we found robust co-occurrence of IgM, IgG, and IgA antibodies reactive to RBD-S in most samples, particularly in post-day 10 samples (**Figure S4**). Pooling all the data revealed that all pre-day 10 RBD-S responses for all isotypes were low.

Around day 10, IgM targeting RBD-S as well as the switched isotypes IgG and IgA simultaneously rose. While RBD-reactive IgM and IgA responses tapered after 3 weeks post-onset (though remained higher than baseline), those for IgG continued to rise to a plateau that was sustained up to 5 weeks after symptoms onset (the most protracted timepoint measured, **Figure 3A**). Similar patterns were obtained for full spike-reactive antibodies (**Figure 3B**). These results suggesting that during acute infection COVID-19 patients undergo a seroconversion across isotypes to SARS-CoV-2 rather than an expansion of pre-existing anti-CoV antibodies.

**Figure 3.**
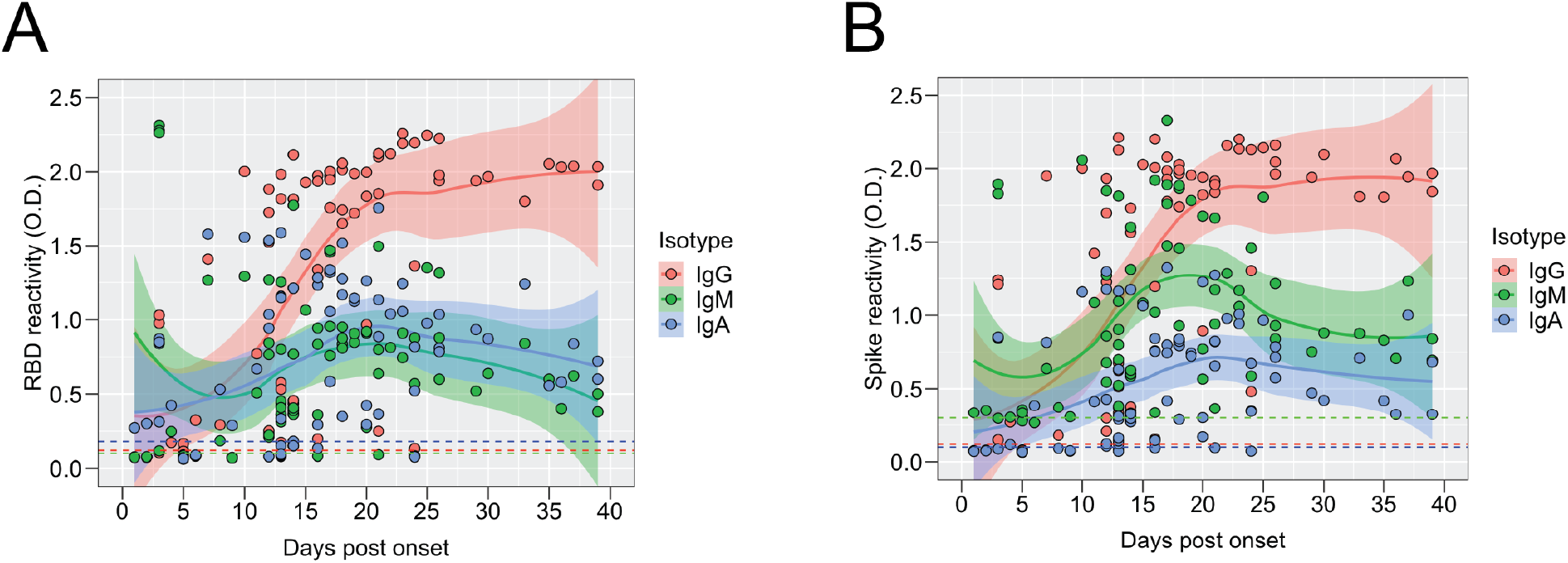
Antibody isotype usage during the response to SARS-CoV-2 RBD-S and spike. **(A)** RBD-S IgM, IgG, and IgA in serum (diluted 1:50) were determined by ELISA and plotted against days post onset of symptoms. LOESS-smoothed lines and 95% confidence intervals are shown for each isotype. (B) **(A)** Spike-reactive IgM, IgG, and IgA in serum (diluted 1:100) were determined by ELISA and plotted against days post onset of symptoms. LOESS-smoothed lines and 95% confidence intervals are shown for each isotype.

Lastly, we assessed anti-RBD-IgG responses by clinical severity. All the patients in this study were hospitalized and 40% of were admitted to the intensive care unit. When we stratified by ICU admission and compared RBS-S IgG levels, we found a trend towards higher levels in those requiring ICU-level care (*P* = 0.09) (**Figure 4A**). Additionally, we observed a significant association between RBD-S IgG and duration of ICU admission (**Figure 4B**). Lastly 7 of 32 (22%) patients succumbed to COVID-19. While a significant difference in the median RBD-S IgG was not observed between survivors and decedents, a smaller range trending towards higher RBD-S reactivity was observed in those patients that died (**Figure 4C**). Although we did not have continuous monitoring of viral load in these patients during hospitalizations it is possible that RBD-S IgG levels reflect ongoing viral replication during more severe disease and in conjunction with other factors may allow for recovery.

**Figure 4.**
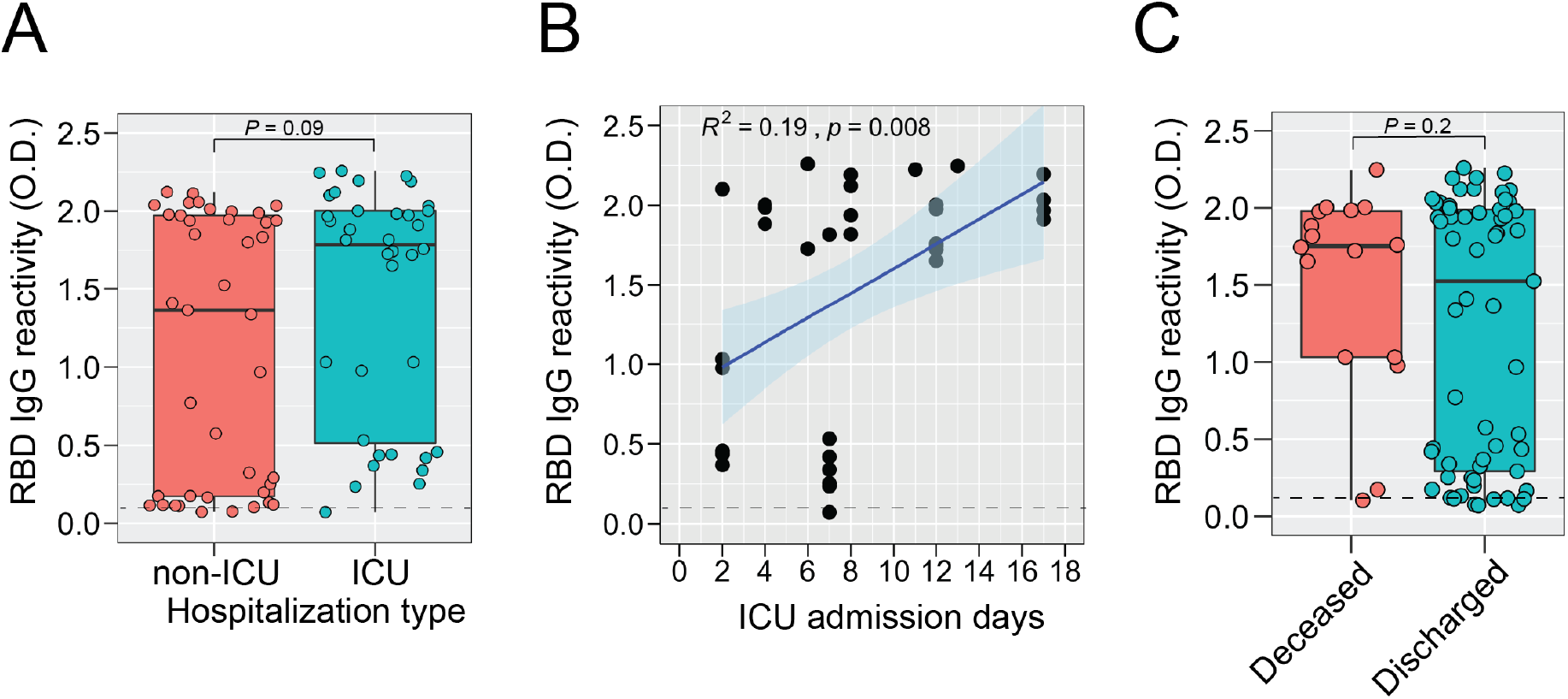
SARS-CoV-2 RBD-S IgG responses during hospitalization. **(A)** RBD-S IgG in patients that were hospitalized in the ICU or not were analyzed by student’s t-test and *P*-value is shown. Boxplots show the median, 95% confidence level and all individual samples. (**B**) For ICU hospitalized patients, all RBD-S IgG values are presented as a function of ICU admission days. Spearman’s Rho coefficient (*R*^*2*^), 95% confidence interval, and *P*-value are shown. (**C**) RBD-S IgG in patients that were deceased or discharged were analyzed by student’s t-test and *P*-value is shown. Boxplots show the median, 95% confidence level, and all individual samples.

Taken together, our results provide the first comprehensive survey of SARS-CoV-2 spike RBD antibodies that accounts for two key risk factors for COVID-19. Neither RBD-S nor S antibodies were significantly different as a function of biological sex. Anti-RBD-S and spike IgG responses were induced across 6 decades of age with robust responses found in several samples from patients ≥ 80 years old. These results also extend kinetic analyses and confirm the paucity of anti-SARS-CoV-2 anti-spike responses in very early blood samples taken prior to day 10 after symptoms onset (17, 24). We also assessed protective anti-spike RBD responses as a function of level of hospital care and disease severity and found that duration of ICU-level care was associated with higher responses, possibly due to an extended period of SARS-CoV-2 replication during severe disease. A limitation of our study is that we only followed symptomatic patients admitted to hospital; it is unclear whether antibody responses differ in asymptomatic or mildly symptomatic patients. We also did not directly assess whether the RBD-specific antibodies we studied were neutralizing at the clonal level, though we did observe a strong association with polyclonal RBD-S IgG responses and SARS-CoV-2 neutralizing activity. This is in agreement with other reports which confirm that RBD-S IgG levels correlate with neutralizing activity and that the RBD of SARS-CoV-2 is a potent target for neutralizing antibodies (16–18, 20, 21, 33). It will be important to determine whether anti-RBD IgA or even IgM antibodies contribute to blocking activity.

## Methods

### COVID-19 samples

Patients were admitted to the University of Vermont Medical Center (UVMMC), situated in a low-density (26-112 persons/km^2^) catchment area with a COVID-19 diagnosis from a PCR-positive swab testing performed within a CLIA-certified clinical laboratory. University of Vermont Institutional Review Board approval was granted under registration STUDY00881. Samples and patient data were obtained under Exemption 4, Waiver of Consent and UVM/UVMMC HIPAA Authorization under 46.116(f)(1)(3), 46.164.512(i)(1)(2). Patient IDs are coded here as “CDDx.001-032”. Deidentified patient (age, sex) and clinical data (COVID-19 diagnosis, dates of symptom onset, hospitalization, intensive care unit admission) were obtained from the electronic health record.

### RBD-S and spike antigen preparations

pCAGGS plasmids containing hexahistidine-tagged SARS-CoV-2 spike glycoprotein receptor binding domain (RBD-S) and trimerized SARS-CoV-2 (15, 23) were obtained as Whatman spots from Florian Krammer (Mt. Sinai School of Medicine), and transformed into *E*.*coli* to make plasmid stocks. We sequence verified these using pcaggs-F (5’-GTTCGGCTTCTGGCGTGT-3’) and pcaggs-R (5’-TATGTCCTTCCGAGTGAGAG-3’). Plasmids were then transfected into Expi293F cells (Gibco #A14527) and protein was purified by Ni-NTA agarose resin (Qiagen #30230) as described (23). Protein was quantified using bovine serum albumin as a standard (Sigma A4505, Cohn Faction V) and Bradford reagent (Bio-Rad, 5000006). Protein was run on denaturing 4–20% recast protein gels (Bio-Rad 4561094) and visualized by Coomassie blue staining with a 10-190 kDa protein ladder (Invitrogen 10748-010).

Spike Glycoprotein Receptor Binding Domain (RBD) from SARS-CoV-2, Wuhan-Hu-1, was also used as a positive control during assay set up and this reagent was produced in HEK293T cells under HHSN272201400008C and obtained through BEI Resources, NIAID, NIH: Spike Glycoprotein Receptor Binding Domain (RBD) from SARS-Related Coronavirus 2, Wuhan-Hu-1, Recombinant from NR-52306.

### Preparation of CR3022 monoclonal antibody

CR3022 is a SARS-CoV S-specific antibody originally isolated by single chain variable region phage display and then cloned as an IgG1/kappa monoclonal human IgG1/κ (13). We received CR3022 heavy chain (HC) and light chain (LC) cloned into pFUSEss-CHIg-hG1 and pFUSE2ss-CLIg-hK, respectively (Invivogen) from Florian Krammer spotted on filter paper. We resuspended spots in 100 µL TE and transformed 20 µL E. coli (NEB C2987H) with 1 µL followed by growth in the presence of Zeocin (25 µg/mL, Invivogen, for CR3022-HC) and blasticidin (100 µg/mL, Invivogen for CR3022 LC). Midi-preps were then sequenced confirmed CR3022HC (Genbank DQ168569) and LC (Genbank DQ168570) with primer HTLV-5’UTR (forward) 5’-GCTTGCTCAACTCTACGTC-3’ and CR3022-HC in the reverse direction by primer Fc (reverse): 5’CTCACGTCCACCACCACGCA-3’. Recombinant CR3022 was expressed in 293A cells (Invitrogen) by polyethyleneimine (Polysciences Inc.) transfection of 9 µg each of CR3022-HC and LC, culture for 7 days, and protein A agarose bead purification as described (34). IgG was quantified by sandwich ELISA with anti-human IgG (Jackson Immunoresearch 109-005-008) as capture and horseradish peroxidase-conjugated anti-human IgG (Jackson Immunoresearch, 109-005-008) as detection Ab with known human serum as a standard.

### Clinical RBD-S and S IgG ELISA testing

For IgG against RBD-S from SARS-CoV-2 we followed Stadlbauer et al (23) and the Emergency Use Authorization granted to MSSM by the Food and Drug Administration on 4/15/2020 (https://www.fda.gov/media/137029/download). Briefly, for RBD-S IgG levels 96-well plates were coated with 100 ng/well of purified RBD-S and then blocked with 3% milk in phosphate-buffered saline (PBS) containing 0.1% Tween-20 (T). Heat-inactivated (56°C for 1 hr) serum samples were diluted 1:5 in PBS, and 20 µL of this was added to 180 µL of dilution buffer (PBS T + 1% milk) in each well for 1:50 final dilution of sample. 100 µL of sample is then added to each well and after 1 hr incubation at room temperature and washing with PBS-T using a Biotek ELx-405 Select CW (Biotek, Winooski, VT), IgG was detected with alkaline phosphatase-conjugated cross-adsorbed anti-human IgG (Sigma SAB3701277, diluted 1:2,500 in blocking buffer), washing, and addition of p-nitrophenylphosphate (Sigma N2770) substrate. The colorimetric reaction (optical density at 405 nm) was detected with a Cytation 3 (Biotek, Winooski, VT). Two negative control samples of pre-2019 serum were used on each plate and the average + three standard deviations above the mean were used as the assay cutoff for positivity.

For S detection heat-inactivated serum samples were diluted 1:5 in PBS and then 20 µL was added to 180 µL of dilution buffer in the starting well (for a final 1:100 starting dilution) then serially diluted 1:3 to an endpoint dilution of 1:8,100. IgG detection was performed as described above with 100 µL of 1:100 sample. Endpoint titer was defined as the last dilution at which the signal was above the cutoff (defined as was done for RBD-S above). spike area under the curve (AUC) was calculated in Prism 8.4.3 (Graphpad Inc) from the OD_405nm_ values from all six dilutions and using the negative control cutoff values as the baseline.

### Testing for RBD-S and S IgM and IgA in clinical samples by ELISA

Samples were handled as above for IgG except that the detection steps used alkaline phosphatase-conjugated anti-human IgM (Sigma A3437, diluted 1:1,000 in blocking buffer) or IgA (Sigma A3400, diluted 1:1,000 in blocking buffer).

### SARS-CoV-2 microneutralization assay

All experiments featuring infectious SARS-CoV-2 were conducted at the UVM BSL-3 facility under an approved Institutional Biosafety protocol. SARS-CoV-2 strain 2019-nCoV/USA_USA-WA1/2020 (WA1) was generously provided by Kenneth Plante and the World Reference Center for Emerging Viruses and Arboviruses (WRCEVA) at the University of Texas Medical Branch and propagated in African green monkey kidney cells (Vero E6) that were kindly provided by J.L Whitton. Vero E6 cells were maintained in complete Dulbecco’s Modified Eagle Medium (cDMEM) (11965–092) containing 10% fetal bovine serum (FBS) (16140–071), 1% HEPES Buffer Solution (15630–130), and 1% penicillin-streptomycin (15140–122) purchased from Thermo Fisher Scientific (Carlsbad, CA). Cells were grown in a humidified incubator at 37°C with 5% CO_2_. To assess the neutralization capacity of patient sera against authentic SARS-CoV-2, we conducted a focus reduction neutralization test (FRNT). Each serum sample was heat inactivated via incubation at 56 °C for 1 h. Samples were then diluted serially in 25 µL of cDMEM, mixed with an equal volume of cDMEM containing 175 focus forming units (FFU) of SARS-CoV-2, and then incubated for 60 minutes at 37°C. Each serum sample was tested for neutralization at an initial dilution of 1:50 and then serially at 1:2 dilutions until reaching an endpoint of 1:3,200. The media from confluent Vero E6 cell monolayers in 96-well white polystyrene microplates (07-200-628, Thermo Fisher Scientific) was removed and 50 µL of each antibody-virus mixture was inoculated onto the cells and incubated at 37°C in a 5% CO_2_ incubator for 60 minutes, after which the wells were overlaid with 1.2% methylcellulose in cDMEM and incubated at 37°C in a 5% CO_2_ incubator for 24 h. Infected cells were fixed in 25% formaldehyde in 3X phosphate buffered saline (PBS). Cells were permeabilized with 0.1% 100X Triton in 1X PBS for 15 minutes and then incubated with a primary, cross-reactive rabbit anti-SARS-CoV N monoclonal antibody (40143-R001, Sinobiological) (1:20,000) followed by a peroxidase-labeled goat anti-rabbit antibody (5220-0336, SeraCare) (1:2,000) and then the peroxidase substrate (5510-0030, SeraCare). Images of the wells were captured using a Zeiss AxioCam MRC Imager.M1 microscope and viral foci were quantified manually. Focus counts were normalized to virus only control wells. FRNT_50_ determinations were made using a non-linear regression curve fit (log[inhibitor] vs. normalized response – variable slope) in GraphPad Prism.

### Graphics and Statistical testing

All statistics and graphics were performed using R version 3.6.1 using standard packages or GraphPad Prism 8.4.3. Non-parametric LOESS (LOcal regrESSion) was used for smoothing.

## Data Availability

All data are presented in the manuscript.

## Acknowledgements

We thank all health care workers and laboratory personnel who contributed to treatment and diagnosis of these and other COVID-19 patients. We thank the clinical research staff at the University of Vermont (UVM) Medical Center Pathology and Laboratory Medicine and the Vaccine Testing Center. We also thank the UVM Research Protections Office, Institutional Review Board, and Institutional Biosafety Committee for rapid turnaround of COVID-19-related projects. The funders had no role in study design, data collection and analysis, decision to publish, or preparation of the manuscript.

## Author contributions

SAD, JWC, and JWB conceived and designed the project. NRG, ANW, CAS, BDM, and SAD performed experiments. AKM, DG, and JWC provided samples. EAB, JWB, MEP, KKP, BDK, RDS, GA, and SAD provided resources and/or key project input. SAD wrote the manuscript with input from all authors. Supervision: SAD, JWC, and JWB.

